# Real-world Effectiveness of Casirivimab and Imdevimab in Patients With COVID-19 in the Ambulatory Setting: An Analysis of Two Large US National Claims Databases

**DOI:** 10.1101/2022.02.28.22270796

**Authors:** Wenhui Wei, Dana Murdock, Jessica J. Jalbert, Vera Mastey, Robert J. Sanchez, Boaz Hirshberg, David M. Weinreich, Mohamed Hussein

**Affiliations:** Regeneron Pharmaceuticals, Inc., Tarrytown, New York, NY

**Author notes:** Corresponding to: Wenhui Wei, PhD, Alternate corresponding author: Mohamed Hussein, PhD.

## Abstract

**Background**

In a phase III clinical trial, casirivimab and imdevimab (CAS+IMD) reduced the composite endpoint of COVID-19-related hospitalizations or all-cause mortality in outpatients at risk of severe disease. This study assessed real-world effectiveness of CAS+IMD.

**Methods**

Data from Optum^®^ Clinformatics^®^ Data Mart (CDM) and IQVIA Pharmetrics Plus (PMTX+) were used to identify patients diagnosed with COVID-19 in ambulatory settings between December 2020 and March 2021 (PMTX+) and June 2021 (CDM), and either treated with CAS+IMD or untreated but treatment-eligible under Emergency Use Authorization. CAS+IMD-treated patients were matched to untreated patients and followed up to 30 days for the outcome of all-cause mortality or COVID-19-related hospitalizations (CDM) and COVID-19-related hospitalizations (PMTX+). Kaplan-Meier estimators were used to calculate outcome risks; Cox proportional-hazard models estimated adjusted hazard ratios (aHR) with 95% confidence intervals (CI).

**Results**

For CDM, 1116 CAS+IMD-treated patients were matched to 5294 untreated patients; for PMTX+, 3280 CAS+IMD-treated patients were matched to 16,284 untreated patients. The 30-day outcome risk was 2.1% and 5.3% in treated and untreated cohorts, respectively (CDM), and the 30-day risk of COVID-19-related hospitalization was 1.9% and 4.8%, respectively (PMTX+); translating to a 61% lower adjusted outcome risk (CDM aHR 0.39 (95% CI 0.26–0.60; PMTX+ aHR 0.39 (95% CI 0.30–0.51). The benefit of treatment was maintained across multiple subgroups of high-risk patients; earlier intervention was associated with improved outcomes.

**Conclusions**

This real-world study further supports randomized clinical trial findings that treatment with CAS+IMD reduces the risk of hospitalization and mortality in patients infected with susceptible variants.

---

Vaccines, in combination with non-pharmaceutical interventions including masking and social distancing, are the mainstay in global efforts to control the ongoing coronavirus 2019 (COVID-19) pandemic caused by severe acute respiratory syndrome coronavirus 2 (SARS-CoV-2). Despite the availability of vaccines, COVID-19-related hospitalizations and mortality remain high in the US and other industrialized nations.

In a clinical trial, intravenous casirivimab and imdevimab (CAS+IMD; REGEN-COV^®^) was shown to reduce all-cause mortality or COVID-19-related hospitalization by 71% among patients with mild-to-moderate disease diagnosed and treated in the ambulatory setting [1]. In November 2020, the combination therapy was authorized under an Emergency Use Authorization (EUA) in the US for the treatment of mild-to-moderate COVID-19 patients who are at risk for severe disease [2]. In June 2021, the subcutaneous administration of CAS+IMD received EUA as an alternative when intravenous infusion is not feasible and would delay treatment. In January 2022, the FDA amended the EUA of CAS+IMD to exclude its use in geographic regions where, based on available information including variant susceptibility and regional variant frequency, infection or exposure is likely due to a variant such as Omicron (B.1.1.529); because of markedly reduced neutralization activity, CAS+IMD is not expected to be active against Omicron [3].

While multiple studies have reported on the real-world effectiveness of CAS+IMD [4–16], most only included a small number of CAS+IMD-treated patients, generally evaluating the early experience with monoclonal antibodies (mAbs), were single-center, and did not have sufficient sample size to assess effectiveness of CAS+IMD across various patient subgroups. Studies leveraging large databases with sufficient counts to assess the effectiveness of CAS+IMD in subgroups of patients at higher risk of poor outcomes are needed. The objective of this study was to compare the effectiveness of CAS+IMD versus no COVID-19 mAb treatment on 30-day all-cause mortality or COVID-19-related hospitalization among patients diagnosed with COVID-19 in the ambulatory setting overall and among subgroups of patients at high-risk of poor COVID-19 outcomes.

## METHODS

This retrospective cohort study was conducted in the US using data from the Optum Clinformatics Data Mart (CDM) and the IQVIA Pharmetrics Plus (PMTX+) databases. Since both databases contain de-identified data and are fully compliant with the Health Insurance Portability and Accountability Act, institutional review board/ethics committee approval was not required. Study design and statistical analyses were described and specified *a priori* in a study protocol.

### Data Sources

The CDM database contains administrative health claims for 68 million members enrolled in large commercial and Medicare Advantage geographically diverse health plans affiliated with Optum since 2007. Information collected includes demographics, inpatient and outpatient claims, outpatient pharmacy claims, and mortality data. The PMTX+ is a national claims database of commercial health plans that includes the records of approximately 190 million patients in all 50 states since 2006. It consists of adjudicated inpatient and outpatient medical claims and outpatient pharmacy claims with month-by-month information on enrolled patients. However, in contrast to CDM, information was not available on mortality during the study period.

### Study Population

Patients with either a COVID-19 diagnosis (International Classification of Diseases [ICD]-10: U07.1) or a positive COVID-19 virus test (CDM only) in the outpatient setting between December 1, 2020, and March 31, 2021 (PMTX+) or June 30, 2021 (CDM) were eligible for inclusion. Patients who were hospitalized as a result of the encounter were excluded. Among eligible patients, we identified those who received CAS+IMD (date of treatment was defined as the index date) and a group of patients who were not treated with any COVID-19 mAb but were eligible to receive CAS+IMD under the EUA. These untreated patients were assigned a random index date drawn from the distribution between days from COVID-19 diagnosis and treatment for CAS+IMD-treated patients. Additional inclusion criteria were continuous healthcare plan enrollment for at least 6 months pre-index (ie, baseline period); ≥12 years of age at index; and diagnosis for COVID-19 or positive test in the 10 days prior to or on the index date (days 0 to –10) but no diagnosis in the previous 30-day period (days –11 to –40) in order to capture the first manifestation of COVID-19 infection rather than complication or follow-up care from a previous episode; and have a valid date of death (CDM only). Patients were excluded if they received COVID-19 mAbs during baseline or multiple COVID-19 mAbs on the index date; or died or were hospitalized between the COVID-19 diagnosis and the index date, inclusive.

### Outcomes

Outcomes were the composite endpoint 30-day all-cause mortality or COVID-19-related hospitalizations for CDM, and 30-day COVID-19-related hospitalizations for PMTX+. A COVID-19-related-hospitalization was defined as hospitalization with a COVID-19 diagnosis as the primary or admitting diagnosis. In CDM, mortality information is derived from the Centers for Medicare and Medicaid Services (CMS), the Social Security Administration (SSA) Death Master File, facility claims indicating that the patient was deceased when discharged, and member coverage information stating the reason for coverage discontinuation was death. PMTX+ does not contain patient mortality data.

Patients were followed from the index date until occurrence of the outcome or censored at the end of 30-day risk period, end of study period (March 31, 2021 for PMTX+, June 30, 2021 for CDM), healthcare plan disenrollment, or use of another COVID-19 mAb, whichever came first.

### Study Variables

Baseline characteristics included demographic variables of age, sex, race/ethnicity, and state. Additional variables included location of the first COVID-19 diagnosis (emergency room/urgent care vs not), number of days from diagnosis to index date; index month; and COVID-19 vaccination status identified using ICD-10 codes. The Charlson Comorbidity Index (CCI) score [17] was determined using ICD-10 diagnosis codes to identify the presence of CCI comorbidities during the baseline period. We also used ICD-10 codes to identify body mass index (BMI), categorized as normal, overweight, obese, severely/morbidly obese; comorbid conditions such as cardiovascular disease (myocardial infarction, hypertension, atrial fibrillation, heart failure, ischemic/hemorrhagic stroke, or venous thromboembolism), chronic respiratory disease (asthma, chronic obstructive pulmonary disease, obstructive sleep apnea, pulmonary fibrosis, cystic fibrosis), cancer, and diabetes (type 1 or 2). Hospitalizations or emergency room/urgent care visits during the baseline period were also captured.

Risk factors based on EUA criteria at the time of the initial CAS+IMD authorization were identified using ICD-10 diagnosis codes, procedure codes, and drug dispensation/administration codes over the baseline period. The EUA criteria included ≥65 years of age; 12–17 years of age and BMI ≥85th percentile for their age and sex based on Centers for Disease Control and Prevention growth charts [18]; BMI >25 kg/m^2^; pregnancy; chronic kidney disease; diabetes; chronic lung disease; immunosuppressive disease; history of immunosuppressive treatment; cardiovascular disease, hypertension, or congenital heart disease; sickle cell disease; neurodevelopmental disorders; and use of medical-related technological dependence [19].

### Statistical Analysis

#### Matching

To control for confounding, untreated EUA-eligible patients were matched to those treated with CAS+IMD without replacement, using exact and probabilistic methods. Propensity score (PS) models predicting the probability of CAS+IMD versus no treatment were derived by including demographic characteristics, individual EUA criteria, CCI score, baseline period hospitalization, days from diagnosis to index date, and emergency room as the ambulatory setting of the diagnosis. CAS+IMD-treated patients were then matched to up to 5 EUA-eligible untreated patients on state, index month, BMI group, and within a caliper width of 0.02 of the PS scale. A variable matching ratio was used to balance between minimizing bias and maximizing the matched sample size to strengthen the statistical power and improve generalizability [20]. Standardized mean differences (SMD) were used to assess balance between treatment groups; SMD >10 indicates imbalance.

#### Primary Analysis

Descriptive analyses of baseline characteristics included means and medians with their standard deviations and interquartile ranges, respectively, for continuous variables, and number and frequency for categorical variables. Kaplan-Meier (KM) time-to-event analyses were conducted on the matched cohorts [21] to provide a nonparametric estimate of the risk of 30-day outcomes. The Hall-Wellner method was used to construct a 95% confidence band across the entire KM survival curve [22]. Log-rank tests were used to compare survival distributions between the 2 cohorts. Cox proportional-hazards models were used to estimate adjusted hazard ratios (aHR) with their 95% confidence intervals (CI) by fitting the model to the matched pairs.

#### Subgroups

We estimated the effectiveness of CAS+IMD versus untreated EUA-eligible patients across subgroups of patients, including: 1) age (<55, 55–64, and ≥65 years), 2) elevated risk defined by any one of the following: ≥65 years old, or 55–64 years of age *and* having at least one of the following: BMI ≥35 kg/m^2^, type 2 diabetes, chronic kidney disease, or chronic obstructive pulmonary disease, 3) number of EUA risk factors (0–2 and ≥3 risk factors), 4) cancer diagnosis or receiving chemotherapy during baseline, 5) B-cell deficiencies defined as over the baseline period having ≥1 inpatient claim or ≥2 outpatient claims dated ≥30 days apart with diagnosis for primary or secondary B-cell deficiency or exposure to ≥1 medication that may induce B-cell deficiency (**Supplementary Table 1**). To investigate the effect of delayed treatment among patients treated with CAS+IMD, outcomes were stratified by days from initial COVID-19 diagnosis to treatment: 1 day, 2 days, 3–4 days, and ≥5 days. The 30-day outcome risks were estimated for each subgroup using KM estimators, and Cox proportional-hazards models were used to derive aHRs by sub-setting to the subgroup of interest and fitting the model to matched pairs.

**Figure 1.**
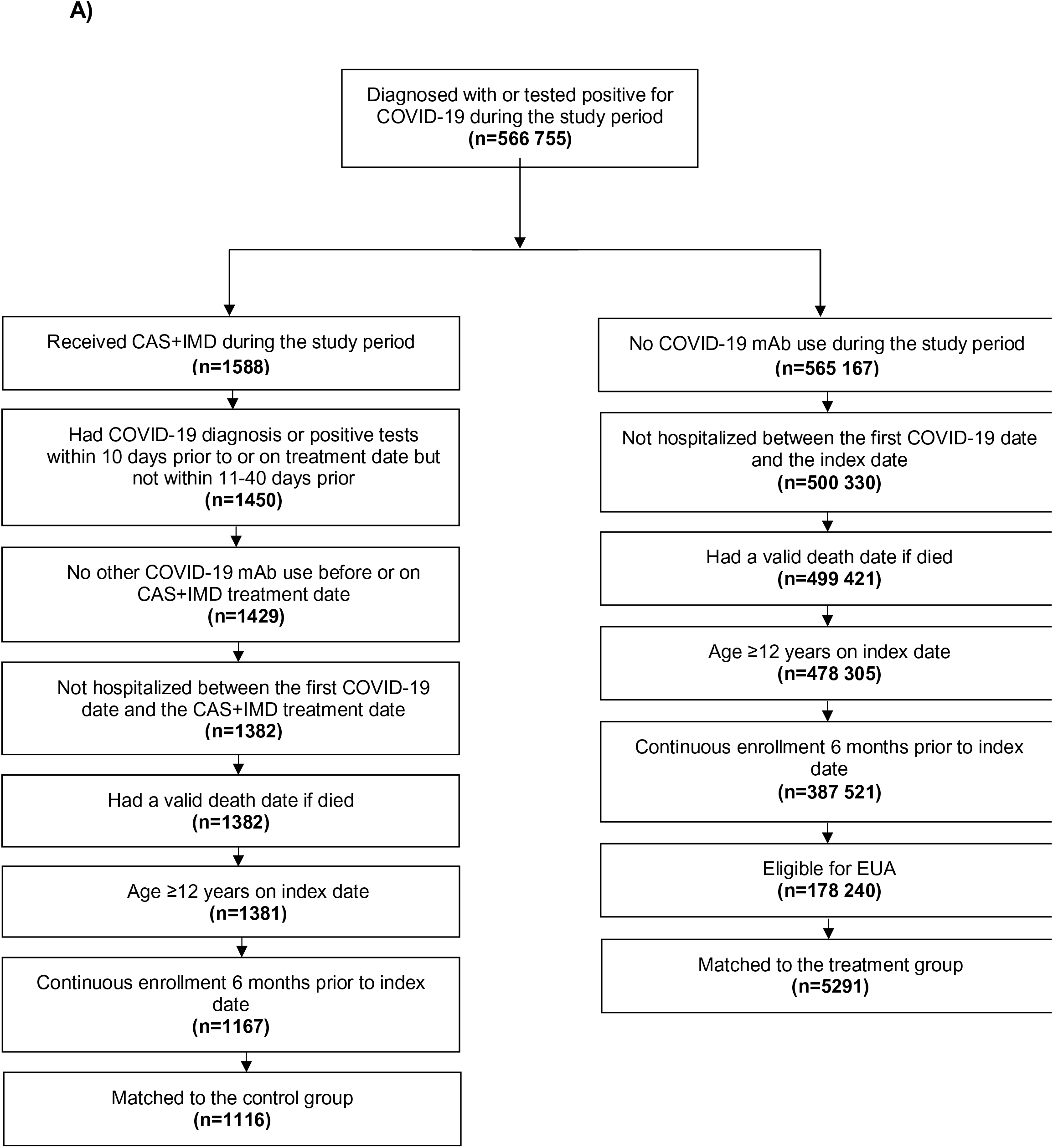
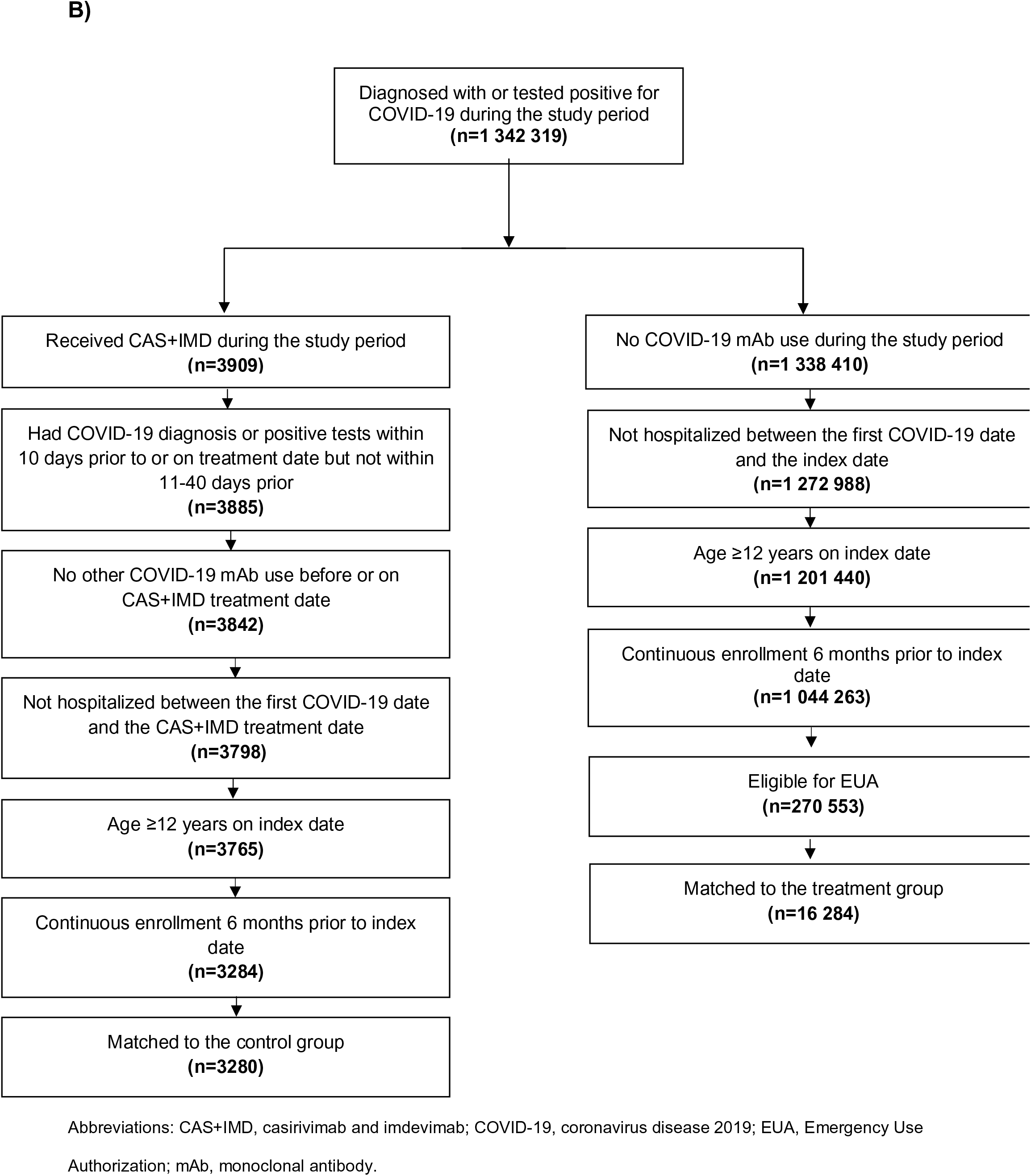
Attrition diagram. Data from (A) Optum CDM database and (B) IQVIA PMTX+ database.

No adjustments were made for multiple comparisons; all *P*-values are nominal with significance set at α = .05.

#### Sensitivity Analyses

Sensitivity analyses included changing the definition of a COVID-19-related hospitalization to a hospitalization with a confirmed COVID-19 diagnosis in the primary position only; changing the definition of a COVID-19-related hospitalization to a hospitalization with a confirmed COVID diagnosis in any position; requiring that treated patients be treated within 7 days of a COVID-19 diagnosis; requiring that the EUA criteria be met for both cohorts; and removal of the COVID-19 diagnosis setting from the propensity score.

The analytic file was created, and all analyses were conducted using Instant Health Data platform (Panalgo, Boston, MA, USA).

## RESULTS

### Patient Populations

In the CDM database, before matching, we identified 1167 patients diagnosed with COVID-19 in ambulatory settings who were treated with CAS+IMD and met all other inclusion criteria and 178 240 untreated EUA-eligible patients (**Figure 1A and Supplementary Table 2**). Of these, 1116 CAS+IMD-treated patients were matched with 5291 of the untreated patients. Demographic characteristics of the matched cohort showed that the treated and untreated groups were primarily Caucasian (∼70%), ∼52% were males, mean age ± standard deviation was 53.0 ± 12.6 and 54.1 ± 12.6 years, respectively, and the greatest representation was from the Southern US (54.4% and 54.8%, respectively) (**Table 1 and Supplementary Table 3**).

**Figure 2.**
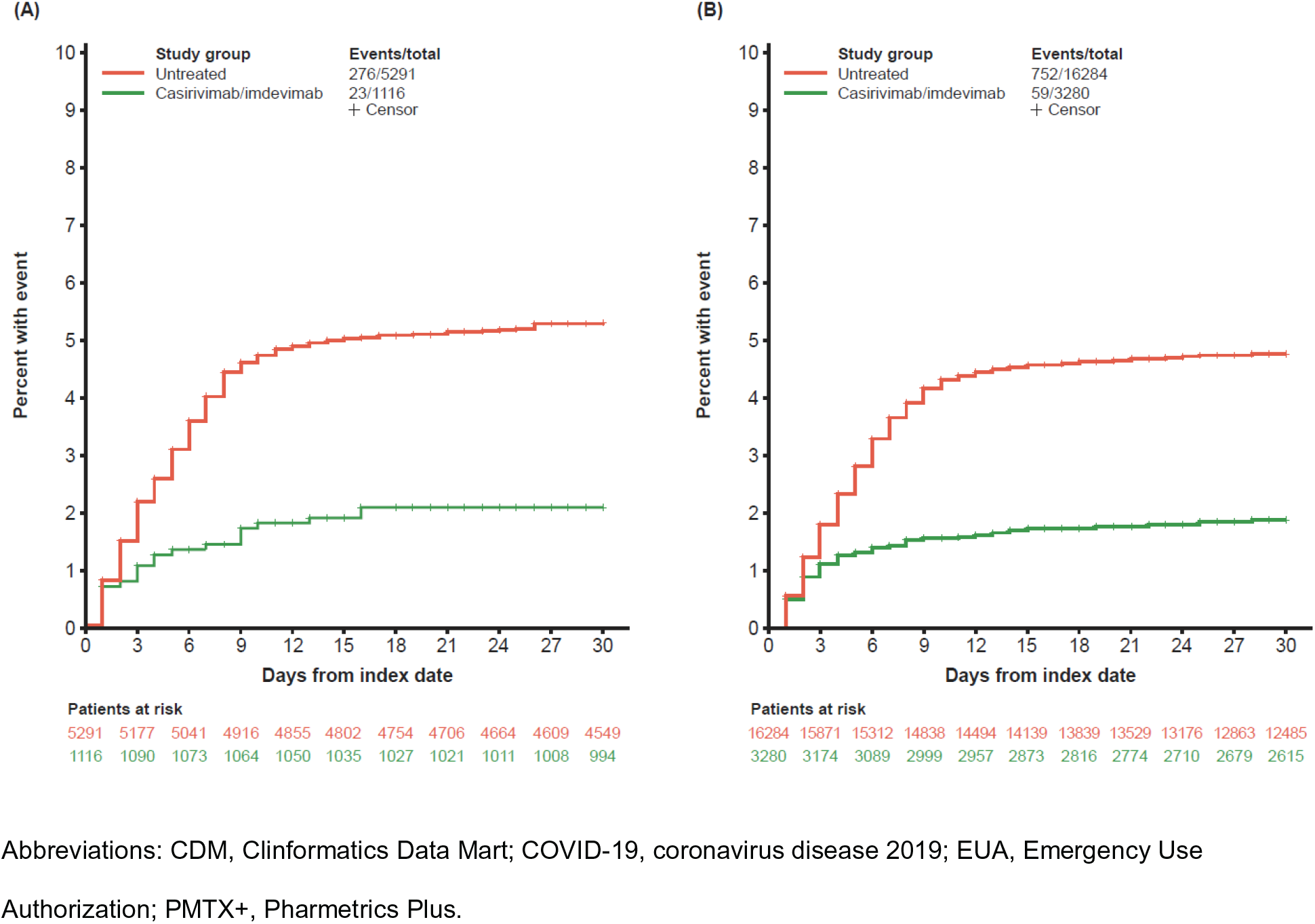
Kaplan-Meier curves of 30-day post-index all-cause mortality and COVID-19-related hospitalization among patients diagnosed with COVID-19 in the outpatient setting who were treated with casirivimab and imdevimab compared with untreated EUA-eligible patients. (A) All-cause mortality or COVID-19-related hospitalization (Optum CDM database) and (B) COVID-19-related hospitalization (IQVIA PMTX+ database).

**Figure 3.**
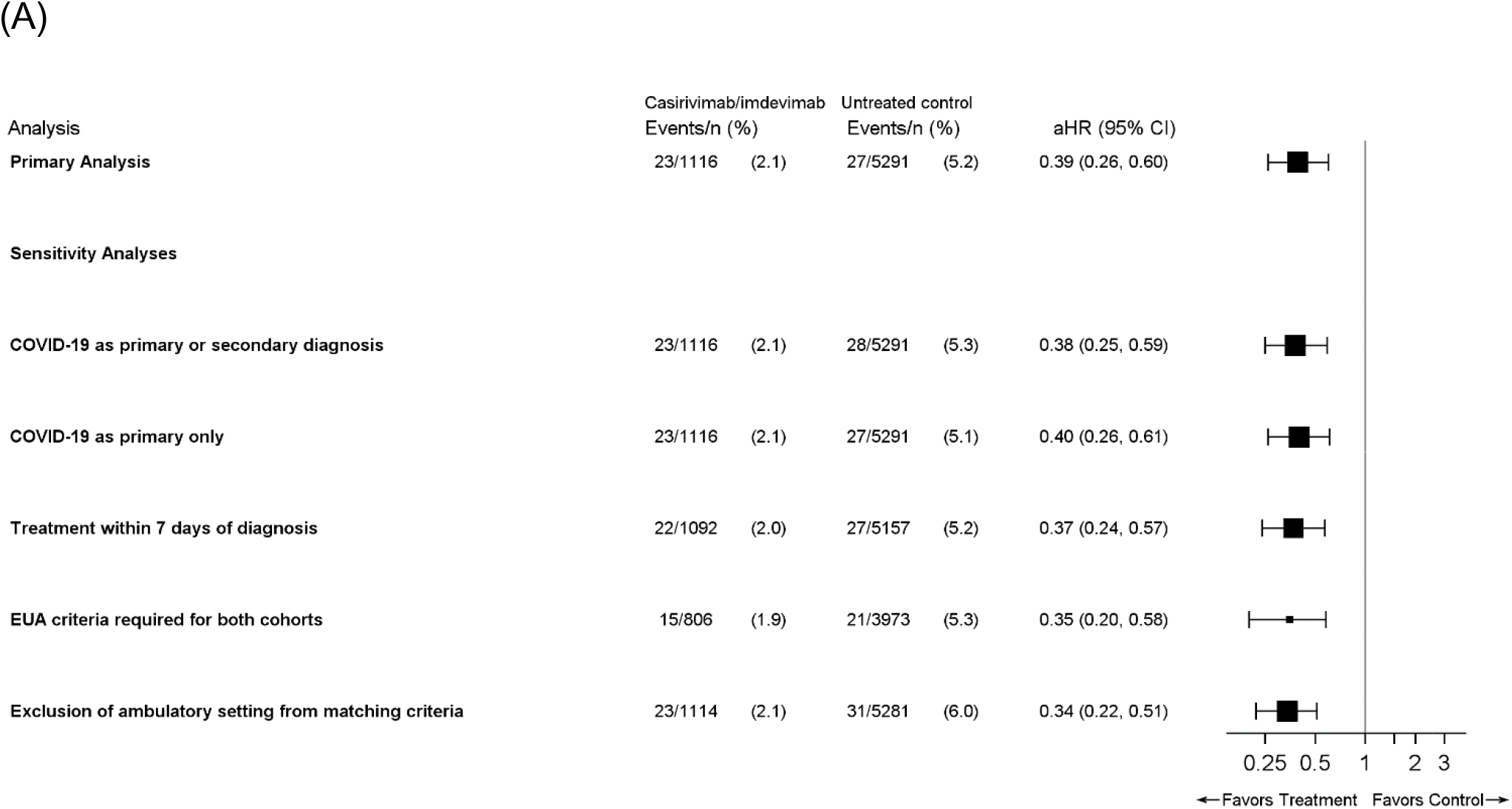
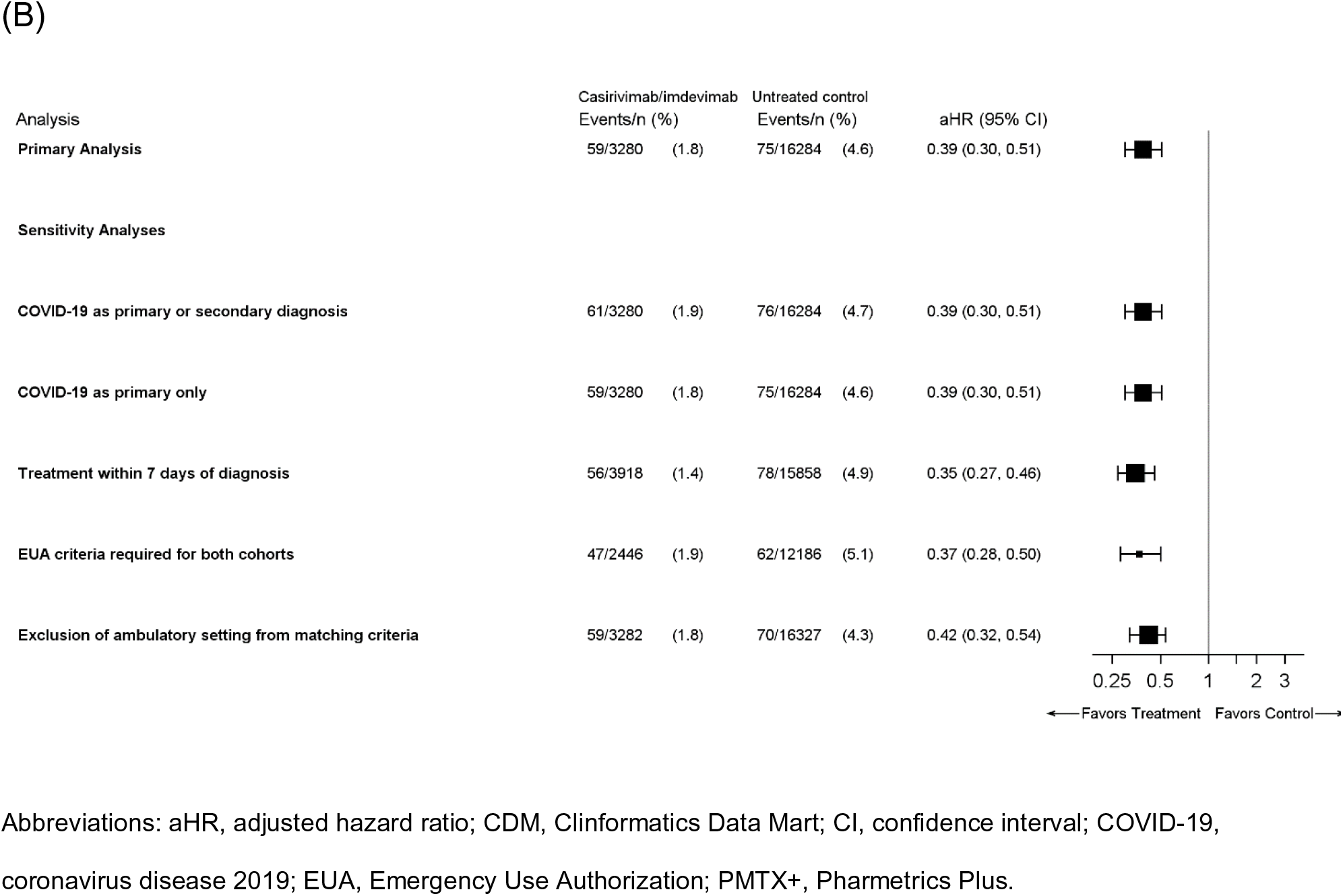
Cox proportional hazards model estimates of aHR with their 95% CIs of 30-day post-index outcomes in matched cohorts. (A) All-cause mortality or COVID-19-related hospitalization (Optum CDM database). (B) COVID-19-related hospitalization (IQVIA PMTX+ database). Square size corresponds to the total sample available for analysis.

**Table 1.** Baseline Characteristics of the Matched Cohorts

| Variable | Optum CDM Database |  |  | IQVIA PMTX+ Database |  |  |
| --- | --- | --- | --- | --- | --- | --- |
|  | CAS+IMD<br>(n = 1116) | No mAb<br>treatment<br>(n = 5291) | SMD | CAS+IMD<br>(n = 3280) | No mAb treatment<br>(n = 16 284) | SMD |
| Age, y |  |  |  |  |  |  |
| Mean ± SD | 53.0 ± 12.6 | 54.1 ± 12.6 | 8.32 | 55.0 ± 11.8 | 55.6 ± 11.8 | 5.23 |
| Median (IQR) | 55 (45–61) | 56 (47–62) |  | 57 (49–62) | 58 (50–63) |  |
| Range | 13–89 | 12–89 |  | 13–85 | 12–85 |  |
| Age group, y |  |  |  |  |  |  |
| 12–17 | 3 (0.3) | 36 (0.7) | 5.99 | 15 (0.5) | 89 (0.6) | 1.26 |
| 18–34 | 88 (7.9) | 370 (7.0) | 3.40 | 199 (6.1) | 923 (5.7) | 1.70 |
| 35–44 | 170 (15.2) | 665 (12.6) | 7.71 | 375 (11.4) | 1563 (9.6) | 5.98 |
| 45–54 | 264 (23.7) | 1150 (21.7) | 4.59 | 685 (20.9) | 3007 (18.5) | 6.09 |
| 55–64 | 447 (40.1) | 2252 (42.6) | 5.10 | 1545 (47.1) | 8433 (51.8) | 9.38 |
| 65–74 | 115 (10.3) | 666 (12.6) | 7.17 | 342 (10.4) | 1714 (10.5) | 0.32 |
| ≥75 | 29 (2.6) | 152 (2.9) | 1.68 | 119 (3.6) | 555 (3.4) | 1.19 |
| Sex |  |  |  |  |  |  |
| Female | 533 (47.8) | 2523 (47.7) | 0.15 | 1623 (49.5) | 7626 (46.8) | 5.31 |
| Male | 583 (52.2) | 2768 (52.3) | 0.15 | 1657 (50.5) | 8658 (53.2) | 5.31 |
| Race/ethnicity |  |  |  |  |  |  |
| Asian | 24 (2.2) | 96 (1.6) | 2.41 | N/A | N/A |  |
| Black | 113 (10.1) | 553 (10.5) | 1.07 | N/A | N/A |  |
| Caucasian | 778 (69.7) | 3712 (70.2) | 0.97 | N/A | N/A |  |
| Hispanic | 140 (12.5) | 632 (11.9) | 1.83 | N/A | N/A |  |
| Missing/unknown | 61 (5.5) | 298 (5.6) | 0.73 | N/A | N/A |  |
| Region |  |  |  |  |  |  |
| Midwest | 265 (23.8) | 1230 (23.3) | 1.18 | 627 (19.1) | 3109 (19.1) | 0.06 |
| Northeast | 129 (11.3) | 607 (11.5) | 0.27 | 433 (13.2) | 2131 (13.1) | 0.34 |
| South | 605 (54.4) | 2900 (54.8) | 1.20 | 2036 (62.1) | 10 148 (62.3) | 0.51 |
| West | 117 (10.5) | 554 (10.5) | 0.04 | 184 (5.6) | 896 (5.5) | 0.47 |
| BMI (kg/m<sup>2</sup>) category<sup>a</sup> |  |  |  |  |  |  |
| Not overweight | 807 (72.3) | 3839 (72.6) | 0.55 | 2587 (78.9) | 12 898 (79.0) | 0.40 |
| Overweight (25 to <30) | 49 (4.4) | 226 (4.3) | 0.59 | 94 (2.9) | 464 (2.8) | 0.14 |
| Obese (30 to <35) | 60 (5.4) | 281 (5.3) | 0.29 | 156 (4.8) | 771 (4.7) | 0.16 |
| Severely obese (35 to <40) | 72 (6.5) | 333 (6.3) | 0.65 | 185 (5.6) | 914 (5.6) | 0.25 |
| Morbidly obese (≥40) | 128 (11.5) | 612 (11.6) | 0.30 | 258 (7.9) | 1280 (7.8) | 0.18 |
| CCI score, mean ± SD | 0.74 ± 1.30 | 0.77 ± 1.29 | 1.95 | 0.83 ± 1.38 | 0.85 ± 1.38 | 1.98 |
| Hospitalization during baseline period | 97 (8.7) | 431 (8.1) | 1.97 | 177 (5.4) | 818 (5.0) | 1.68 |
| Month of index date |  |  |  |  |  |  |
| December 2020 | 131 (11.8) | 654 (12.4) | 1.91 | 528 (16.1) | 2637 (16.2) | 0.26 |
| January 2021 | 272 (24.4) | 1344 (25.4) | 2.38 | 1412 (43.1) | 7060 (43.2) | 0.42 |
| February 2021 | 109 (9.9) | 528 (10.0) | 0.41 | 697 (21.3) | 3452 (21.1) | 0.40 |
| March 2021 | 169 (15.2) | 798 (15.1) | 0.42 | 643 (19.6) | 3178 (19.5) | 0.36 |
| April 2021 | 219 (19.7) | 1017 (19.2) | 0.79 | N/A | N/A |  |
| May 2021 | 122 (11.0) | 546 (10.3) | 2.56 | N/A | N/A |  |
| June 2021 | 92 (8.3) | 404 (7.6) | 1.92 | N/A | N/A |  |
| Time from diagnosis to index date, days |  |  |  |  |  |  |
| Mean ± SD | 3.0 ± 2.1 | 3.0 ± 1.8 | 0.77 | 2.9 ± 2.2 | 2.9 ± 1.8 | 1.56 |
| Median (IQR) | 2 (1–4) | 3 (1–4) |  | 2 (1–4) | 2 (1–4) |  |
| Range | 1–11 | 1–11 |  | 1–11 | 1–11 |  |
| Initial outpatient COVID-19 diagnosis in ER | 26 (2.3) | 177 (3.4) | 5.74 | 509 (15.5) | 2684 (16.5) | 2.63 |
| Vaccinated | 87 (7.8) | 346 (6.5) | 4.87 | 94 (2.9) | 359 (2.2) | 4.21 |
| EUA high risk criteria |  |  |  |  |  |  |
| Age ≥65 years | 144 (12.9) | 818 (15.6) | 7.33 | 461 (14.1) | 2269 (13.9) | 0.35 |
| Children overweight<sup>b</sup> | 1 (0.1) | 9 (0.2) | 2.46 | 2 (0.1) | 7 (<0.1) | 0.79 |
| Overweight<sup>a</sup> | 260 (23.3) | 1226 (23.2) | 0.30 | 599 (18.3) | 2951 (18.1) | 0.36 |
| Pregnancy | 8 (0.7) | 40 (0.8) | 0.46 | 10 (0.3) | 72 (0.4) | 2.25 |
| Chronic kidney disease | 57 (5.1) | 312 (5.9) | 3.46 | 181 (5.5) | 1263 (7.8) | 9.00 |
| Diabetes | 318 (28.5) | 1685 (31.9) | 7.31 | 1028 (31.3) | 5441 (33.4) | 4.43 |
| Immunocompromised | 177 (15.9) | 1048 (19.8) | 10.32 | 441 (13.5) | 2542 (15.6) | 6.15 |
| Cardiovascular disease or hypertension | 591 (53.0) | 2908 (55.0) | 4.02 | 1857 (56.6) | 9608 (59.0) | 4.83 |
| Chronic pulmonary disease | 186 (16.7) | 930 (17.6) | 2.42 | 516(15.7) | 2934(18.0) | 6.11 |
| Sickle cell disease | 1 (0.1) | 7 (0.1) | 1.28 | 3 (0.1) | 18 (0.1) | 0.60 |
| Neurodevelopmental disorders | 254 (22.8) | 1207 (22.8) | 0.13 | 733 (22.4) | 3430 (21.1) | 3.11 |
| Medical-related technological dependence | 237 (21.2) | 1088 (20.6) | 1.66 | 653 (19.9) | 3127 (19.2) | 1.78 |

Data are presented as no. (%) unless otherwise indicated.
Abbreviations: BMI, body mass index; CAS+IMD, casirivimab and imdevimab; CDM, Clinformatics Data Mart; ER, emergency room; EUA, Emergency Use Authorization; IQR, interquartile range; mAb, monoclonal antibody; N/A, not available; PMTX+, Pharmetrics Plus; SD, standard deviation; SMD, standardized mean difference.
<sup>a</sup>Based on diagnoses relating to the BMI categories.
<sup>b</sup>Based on BMI ≥85th percentile for age and sex among those 12–17 years old.

In the PMTX+ database, 3284 CAS+IMD-treated patients and 270 553 untreated EUA-eligible patients were identified before matching (**Figure 1B and Supplementary Table 2**); 3280 CAS+IMD-treated patients were matched to 16 284 untreated patients. Baseline characteristics were generally similar to the CDM database (**Table 1 and Supplementary Table 3**).

The SMDs showed that, overall, matched patients in each dataset were well-balanced (except for time from diagnosis) (**Table 1**). Additionally, patients in CDM and PMTX+ databases were generally similar except for different regional distribution, index month, location of COVID-19 diagnosis, and vaccination status.

Among those treated with CAS+IMD, the mean and median times from diagnosis to index date showed a generally short duration between diagnosis and treatment (≤3 days) (**Table 1**). The prevalence of COVID-19 risk factors was similar between treated patients in the CDM and PMTX+ databases; cardiovascular disease or hypertension had the highest prevalence, 53.0%–59.0%, followed by diabetes, 28.5%–33.4%, respectively (**Table 1**). In CDM and PMTX+, 12.9%–15.6% of patients were ≥65 years of age, and 18.1%–23.3% had an obesity-related diagnosis (**Table 1**).

### Primary Analysis

In CDM, event rates for the composite endpoint of 30-day post-index all-cause mortality/COVID-19-related hospitalizations were 2.1% (95% CI 1.3–3.0) in the CAS+IMD-treated group and 5.3% (95% CI 4.7–5.9) in the untreated group (**Figure 2A**); most events occurred within the first 10 days post-index. No deaths occurred in the treated cohort during the 30-day post-index period, while 27 deaths occurred in the same period among untreated patients (*P* = .015 for the difference in survival probability).

In PMTX+, the 30-day post-index COVID-19-related hospitalizations were 1.9% (95% CI 1.4–2.3) and 4.8% (95% CI 4.4–5.1) in the CAS+IMD-treated and untreated groups, respectively (**Figure 2B**). Most events occurred within 10 days post-index.

Treatment with CAS+IMD was associated with a 61% lower risk when compared to the untreated EUA-eligible patients for the composite endpoint (aHR 0.39, 95% CI 0.26–0.60; **Figure 3A**) in the CDM population and for COVID-19-related hospitalizations (aHR 0.39, 95% CI 0.30–0.51; **Figure 3B**) in the PTMX+ population. Results were robust across sensitivity analyses (**Figure 3**).

### Subgroup Analyses

Compared to the untreated group, CAS+IMD treatment was associated with a consistently lower risk (28%–88%) of the outcomes across subgroups in both databases (**Figure 4**). The largest effect sizes were observed in patients with cancer/receiving chemotherapy (66% and 75% in CDM and PMTX+, respectively) and B-cell deficiencies (88% and 66% in CDM and PMTX+, respectively). In CDM, the estimate did not reach statistical significance within a few subgroups with small sample sizes in the treated group, including those >64 years of age, >3 EUA risk factors, and those with cancer/receiving chemotherapy).

**Figure 4.**
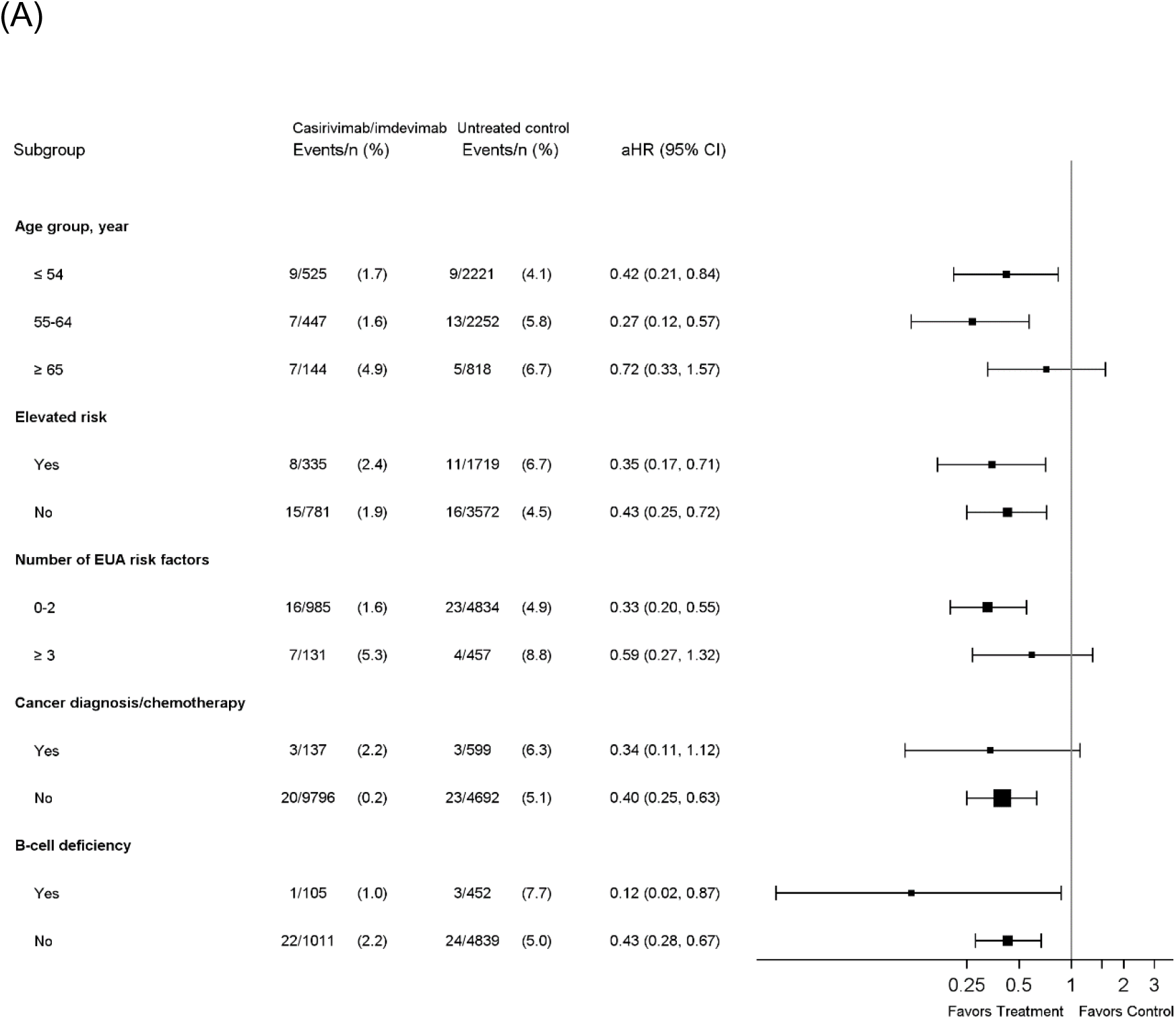
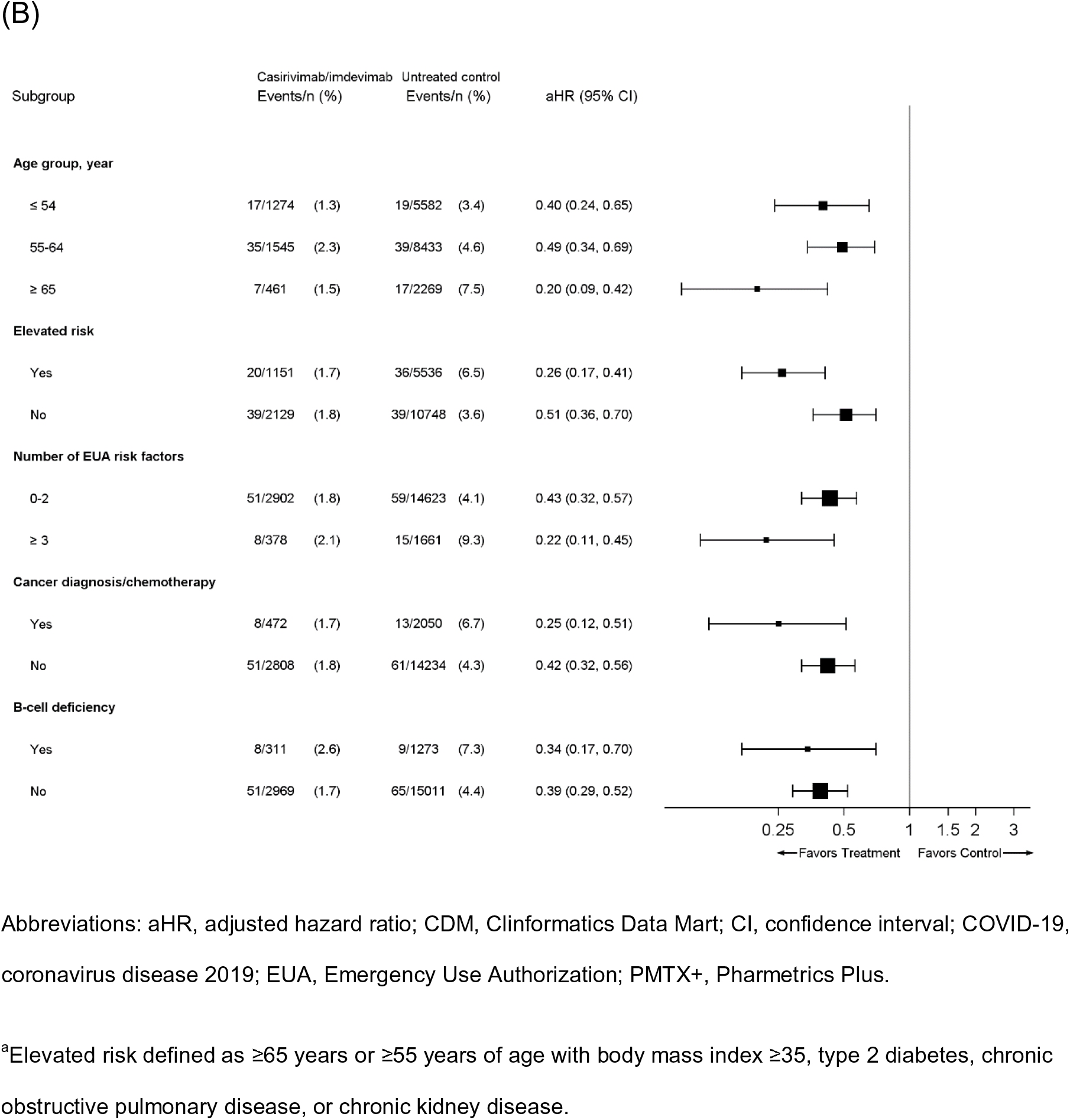
Cox proportional hazards model estimates of aHR with their 95% CIs of 30-day post-index in high risk subgroups. (A) All-cause mortality or COVID-19-related hospitalization (Optum CDM database). (B) COVID-19-related hospitalization (IQVIA PMTX+ database). Square size corresponds to the total sample available for analysis.

**Figure 5.**
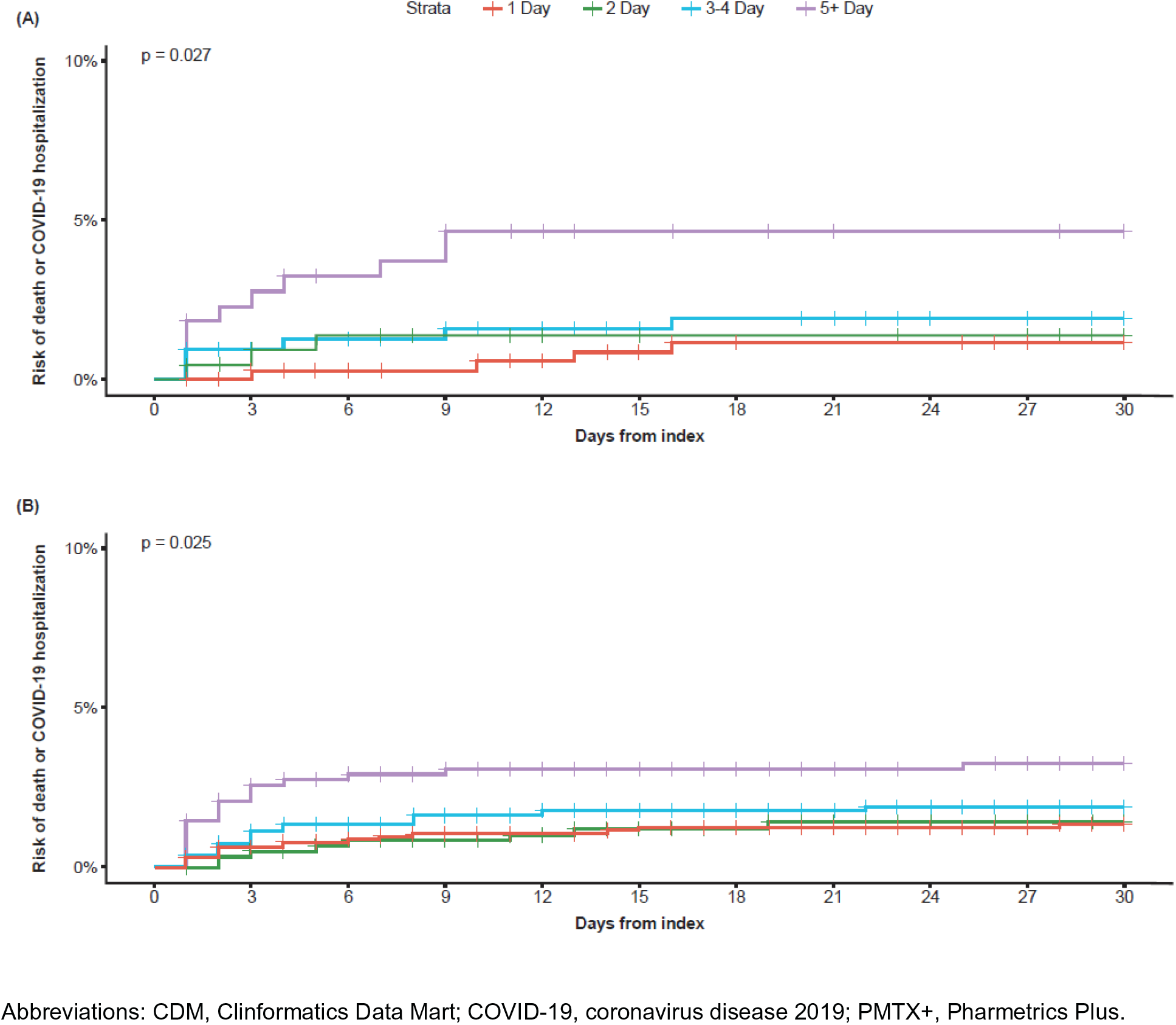
Kaplan-Meier curves of post-index all-cause mortality and COVID-19-related hospitalizations stratified by time from initial COVID-19 diagnosis to index event among treated patients. (A) All-cause mortality or COVID-19-related hospitalization (Optum CDM database) and (B) COVID-19-related hospitalization (IQVIA PMTX+ database). *P*-value derived using the log-rank test.

Among treated patients, earlier treatment after the COVID-19 diagnosis was associated with improved outcomes as assessed by all-cause death/COVID-related hospitalizations in the CDM data ie, 1.2% (95% CI 0.0–2.3) for 1 day and 4.6% (95% CI 1.8–7.4) for ≥5 days (**Figure 4A**). A similar pattern was observed in the PMTX+ data (**Figure 4B**).

## DISCUSSION

This retrospective cohort study suggests that in non-hospitalized patients diagnosed with COVID-19, treatment with CAS+IMD was associated with a significantly lower risk of subsequent 30-day all-cause mortality or COVID-19-related hospitalization relative to no mAb treatment when the variant is susceptible to neutralization by the therapy.

These effects were even more pronounced among patients at elevated risk and those with B-cell deficiencies. Furthermore, these findings were consistent across two large independent databases, affirming the robustness of observed effectiveness of CAS+IMD treatment. These results further support the significant reduction in risk of all-cause mortality or COVID-19 related hospitalization that was previously reported with CAS+IMD in the phase III clinical trial [1].

Evidence from several real-world studies suggests that appropriate treatment with CAS+IMD was associated with an incidence of hospitalizations that may be 50%–90% lower than among untreated patients [4, 5, 8, 9, 16, 23]. The magnitude of effects observed in the current study fell within that range and, additionally, demonstrated greater benefits among those at higher risk. However, it should be noted that there was substantial heterogeneity in populations and outcomes among the real-world studies, including use of a shorter follow-up [9] or more broadly defined endpoints such as all-cause hospitalizations [4, 9, 23] or medically attended visits (ie, hospitalizations and emergency department) [13].

Importantly, treatment with CAS+IMD appeared to have a benefit on mortality in the CDM cohort, as the 30-day estimated risk of death was much lower in the treated than untreated patients. These results further support the public health implications of this therapy, and are consistent with the real-world studies that also suggested lower mortality in patients who received mAb treatment [9, 11, 15, 16], albeit in variants susceptible to such therapy.

The current study also expands on the results of the clinical trial and real-world studies by demonstrating that treatment with CAS+IMD may be especially beneficial for improving outcomes in subgroups of patients at higher risk of severe disease. Among these subgroups are those who are immunocompromised such as patients with a variety of B-cell deficiencies, or those defined as having an elevated risk (ie, ≥65 years or ≥55 years of age with BMI ≥35, type 2 diabetes, chronic obstructive pulmonary disease, or chronic kidney disease).

The results show a clear association between timing of treatment and therapeutic benefit, emphasizing that patients should be treated as soon as possible to receive the maximal benefit of treatment. These results also highlight the need for rapid outpatient diagnosis and subsequent early initiation of treatment for those who meet the EUA criteria.

Potential limitations of this analysis include that claims data do not capture COVID-19 disease severity (eg, viral load) or symptom data, including time of symptom onset, which are important predictors of hospitalization and mortality and may be used by clinicians to inform treatment decisions. Since the study was non-randomized, this limitation may result in residual confounding due to channeling bias; some patients may not have been treated because they may have had milder disease than those who were treated, potentially leading to underestimation of CAS+IMD effectiveness. Potentially important confounders such as BMI and COVID-19 vaccination are not well captured in claims data; assuming patients with higher BMI and those who are unvaccinated would be more likely to have worse disease and be treated, residual confounding would likely bias results against CAS+IMD. Misclassification may similarly result from use of days from diagnosis to treatment as a proxy for the duration from symptom onset to treatment as, for many patients, COVID-19 symptoms are likely to have started prior to diagnosis. Furthermore, the data were sourced at a time when vaccinations were not routinely available, particularly in the PMTX+ database, thus limiting the generalizability of our findings. Additionally, the current findings may not be generalizable to future variants, such as Omicron, where laboratory studies have found markedly reduced neutralization activity [24, 25]. Finally, the claims data were not able to distinguish between subcutaneous and intravenous formulations of CAS+IMD. Despite these limitations, the design and methods employed in this study, as well as, the robustness of results observed in the sensitivity analyses provide a degree of confidence with the results.

In conclusion, results from this real-world study are consistent with and extend those observed in the randomized clinical trials. This study demonstrated that, relative to untreated EUA-eligible patients, in an environment where the variant is known to be neutralized by CAS+IMD, treatment was associated with a significant 61% reduction in the risk of outcomes (CDM: all-cause mortality/COVID-19-related hospitalization; PMTX+: COVID-19-related hospitalization) among high-risk patients diagnosed with COVID-19 in the outpatient setting. Patients with multiple risk factors may especially benefit from treatment, such as those who are immunocompromised (ie, cancer/chemotherapy or B-cell deficiencies) and those at elevated risk. CAS+IMD was also associated with a low absolute event rate, but these rates increased with delayed treatment, suggesting the importance of early diagnosis and treatment in the ambulatory setting.

### Notes

#### Data sharing

Qualified researchers may request access to study documents (including the clinical study report, study protocol with any amendments, blank case report form, and statistical analysis plan) that support the methods and findings reported in this manuscript. Individual anonymized participant data will be considered for sharing once the indication has been approved by a regulatory body, if there is legal authority to share the data and there is not a reasonable likelihood of participant re-identification. Submit requests to https://vivli.org/.

## Supporting information

BH ICMJE

DM ICMJE

DMW ICMJE

JJJ ICMJE

MH ICMJE

RJS ICMJE

VM ICMJE

WW ICMJE

## Acknowledgments

Medical writing support in the preparation of this publication was provided by E. Jay Bienen, PhD, an independent medical writer, and funded by Regeneron Pharmaceuticals, Inc. The authors also thank Prime for formatting and copyediting, Audrey Garman and Shaila Murji from Panalgo, LLC. for data quality check, and Andreas Kuznik from Regeneron Pharmaceuticals, Inc. for providing input during the study.

---

**Supplementary Table 1.**
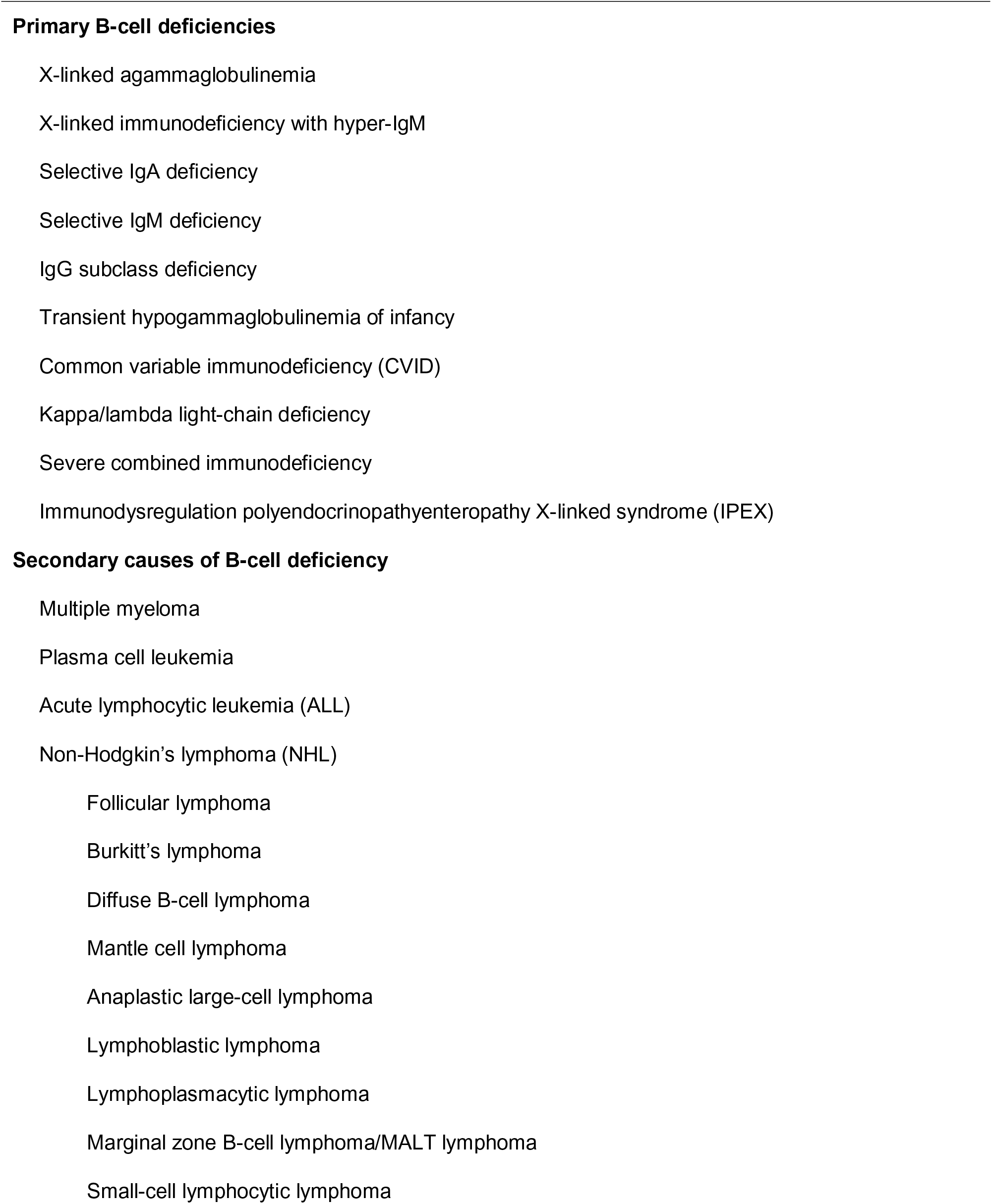
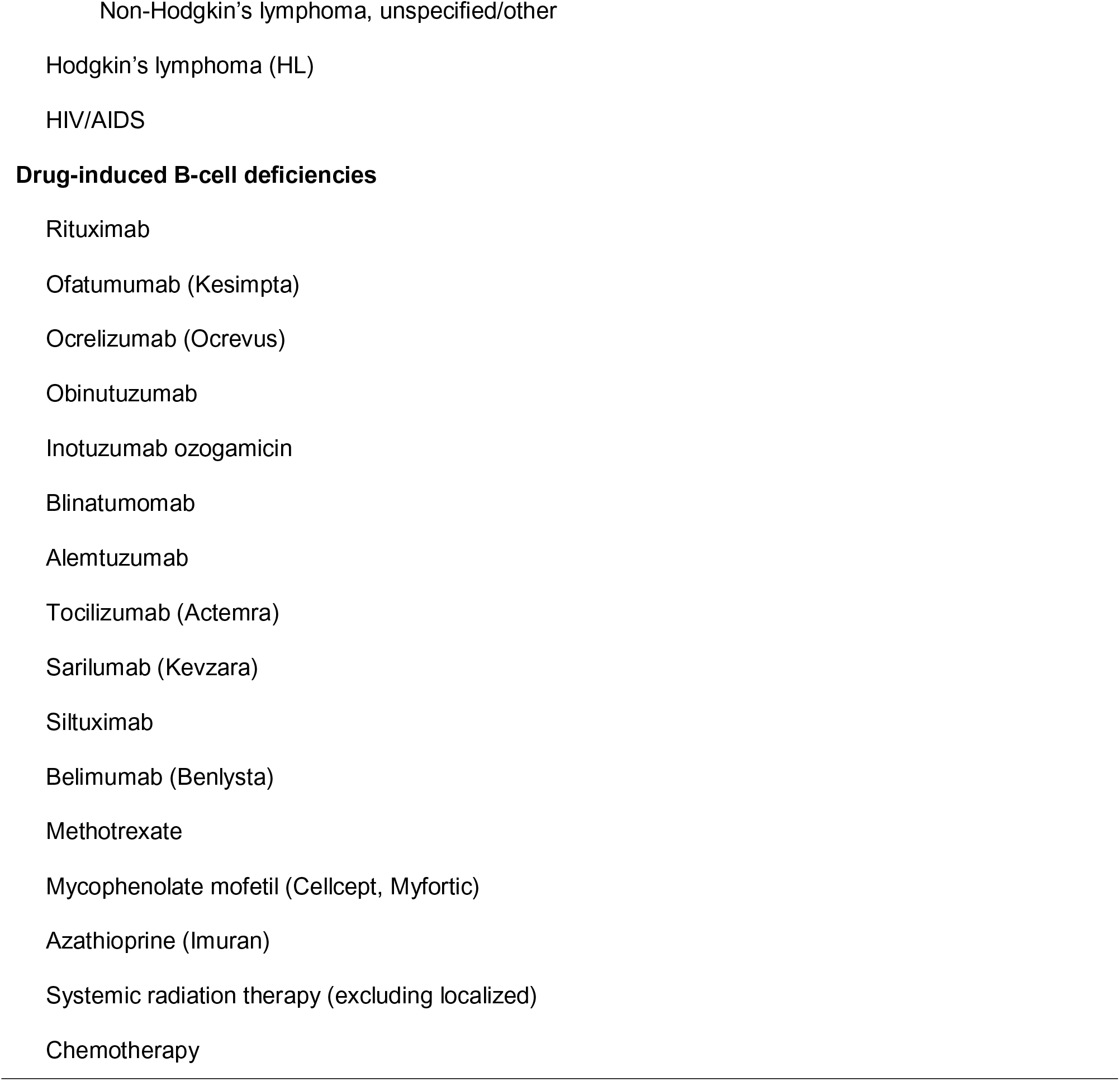
B-cell Deficiencies.

**Supplementary Table 2.** Baseline Characteristics of the Unmatched Cohorts.

| Variable | Optum CDM Database |  | IQVIA PMTX+ Database |  |
| --- | --- | --- | --- | --- |
|  | CAS+IMD<br>(n =1167) | No mAb treatment<br>(n = 178 240) | CAS+IMD<br>(n = 3284) | No mAb treatment<br>(n = 270 553) |
| Age, years |  |  |  |  |
| Mean ± SD | 52.7 ± 12.0 | 65.1 ± 16.3 | 55.0 ± 11.8 | 52.9 ± 16.6 |
| Median (IQR) | 55 (45-61) | 68 (58-76) | 57 (49-62) | 57 (45-63) |
| Range | 18-39 | 12-90 | 13-85 | 12-85 |
| Age group, y |  |  |  |  |
| 12–17 | 3 (0.3) | 5865 (3.3) | 15 (0.5) | 18 852 (7.0) |
| 18–34 | 93 (8.0) | 5775 (3.2) | 199 (6.1) | 20 684 (7.6) |
| 35–44 | 181 (15.5) | 7911 (4.4) | 375 (11.4) | 27 357 (10.1) |
| 45–54 | 296 (25.4) | 13 470 (7.6) | 686 (20.9) | 45 666 (16.9) |
| 55–64 | 450 (38.6) | 31 290 (17.6) | 1548 (47.1) | 103 674 (38.3) |
| 65–74 | 115 (9.9) | 64 859 (36.4) | 342 (10.4) | 36 632 (13.5) |
| ≥75 | 29 (2.5) | 49 070 (27.5) | 119 (3.6) | 17 688 (6.5) |
| Sex |  |  |  |  |
| Female | 547 (46.9) | 101 753 (57.1) | 1624 (49.5) | 146 398 (54.1) |
| Male | 619 (53.1) | 76 485 (42.9) | 1660 (50.5) | 124 155 (45.9) |
| Race/ethnicity |  |  |  |  |
| Asian | 24 (2.1) | 4434 (2.5) | N/A | N/A |
| Black | 117 (10.0) | 20 893 (11.7) | N/A | N/A |
| Caucasian | 819 (70.2) | 113 004 (63.4) | N/A | N/A |
| Hispanic | 143 (12.3) | 25 992 (14.6) | N/A | N/A |
| Missing/unknown | 64 (5.5) | 13 917 (7.8) | N/A | N/A |
| Region |  |  |  |  |
| Midwest | 286 (24.5) | 35 533 (19.9) | 628 (19.1) | 61 026 (22.6) |
| Northeast | 137 (11.8) | 26 488 (14.9) | 433 (13.2) | 33 957 (12.6) |
| South | 620 (53.2) | 87 469 (49.1) | 2039 (62.1) | 149 980 (55.5) |
| West | 123 (10.6) | 28 688 (16.1) | 184 (5.6) | 25 322 (9.4) |
| BMI (kg/m<sup>2</sup>) category<sup>a</sup> |  |  |  |  |
| Not overweight | 833 (71.4) | 132 679 (74.4) | 2588 (78.8) | 196 964 (72.8) |
| Overweight (25 to <30) | 54 (4.6) | 10 557 (5.9) | 94 (2.9) | 10 345 (3.8) |
| Obese (30 to <35) | 70 (6.0) | 11 310 (6.4) | 156 (4.8) | 12 923 (4.8) |
| Severely obese<br>(35 to <40) | 76 (6.5) | 11 271 (6.3) | 187 (5.7) | 23 392 (8.6) |
| Morbidly obese (≥40) | 134 (11.5) | 12 423 (7.0) | 259 (7.9) | 26 929 (10.0) |
| CCI score, mean ± SD | 0.74 (1.31) | 1.18 (1.66) | 0.8 (1.4) | 0.7 (1.2) |
| Hospitalization | 99 (8.5) | 14 918 (8.4) | 177 (5.4) | 16 160 (6.0) |
| Month of index date |  |  |  |  |
| December 2020 | 133 (0.2) | 64 056 (99.8) | 528 (0.5) | 110 879 (99.5) |
| January 2021 | 276 (0.5) | 58 056 (99.5) | 1414 (1.5) | 95 876 (98.5) |
| February 2021 | 118 (0.6) | 21 261 (99.5) | 698 (1.8) | 37 464 (98.2) |
| March 2021 | 177 (1.4) | 12 921 (98.7) | 644 (2.4) | 26 334 (97.6) |
| April 2021 | 225 (2.0) | 11 153 (98.0) | N/A | N/A |
| May 2021 | 133 (1.9) | 6898 (98.1) | N/A | N/A |
| June 2021 | 105 (2.6) | 3895 (97.4) | N/A | N/A |
| Time from diagnosis to index date, days |  |  |  |  |
| Mean ± SD (range) | 2.9 ± 2.1 | 3.1± 1.9 | 2.8± 2.2 | 3.1± 1.9 |
| Median (IQR) | 2 (1-4) | 3 (2-4) | 2 (1-4) | 3 (2-4) |
| Range | 1-11 | 1-11 | 1-11 | 1-11 |
| Initial outpatient COVID-19 diagnosis in ER | 26 (2.2) | 24 009 (13.5) | 511 (15.6) | 27 082 (10.0) |
| Vaccinated | 88 (7.5) | 2575 (1.4) | 94 (2.9) | 3177 (1.2) |
| EUA high risk patients | 966 (82.8) | 178 197 (100) | 2797 (85.2) | 270 383 (99.9) |
| Age ≥65 years | 144 (12.3) | 113 929 (63.9) | 461 (14.0) | 54 320 (20.1) |
| Children overweight<sup>b</sup> | 1 (0.1) | 1198 (0.7) | 2 (0.1) | 2309 (0.9) |
| Overweight<sup>a</sup> | 280 (24.0) | 35,004 (19.6) | 602 (18.3) | 63,244 (23.4) |
| Pregnancy | 8 (0.7) | 616 (0.4) | 10 (0.3) | 1,957 (0.7) |
| Chronic kidney disease | 57 (4.9) | 20 239 (11.4) | 182 (5.5) | 14 233 (5.3) |
| Diabetes | 320 (27.4) | 62 518 (35.1) | 1029 (31.3) | 90 980 (33.6) |
| Immunocompromised | 181 (15.5) | 22 122 (12.4) | 441 (13.4) | 45 924 (17.0) |
| Cardiovascular disease<br>or hypertension | 602 (51.6) | 122 061 (68.5) | 1859 (56.6) | 158 120 (58.4) |
| Chronic pulmonary<br>disease | 198 (17.0) | 33 335 (18.7) | 516(15.7) | 38 860 (14.4) |
| Sickle cell disease | 2 (0.2) | 156 (0.1) | 3 (0.1) | 220 (0.1) |
| Neurodevelopmental<br>disorders | 258 (22.1) | 57 244 (32.1) | 733 (22.3) | 74 600 (27.6) |
| Medical related<br>technological<br>dependence | 245 (21.0) | 44 166 (24.8) | 653 (19.9) | 51 711 (19.1) |

Data are presented as no. (%) unless otherwise indicated.
Abbreviations: BMI, body mass index; CAS+IMD, casirivimab and imdevimab; CDM, Clinformatics Data Mart; ER, emergency room; EUA, Emergency Use Authorization; IQR, interquartile range; mAb, monoclonal antibody; N/A, not available; PMTX+, Pharmetrics Plus; SD, standard deviation; SMD, standardized mean difference.
<sup>a</sup>Based on diagnoses relating to the BMI categories.
<sup>b</sup>Based on BMI ≥85th percentile for age and sex among those 12–17 years old.

**Supplementary Table 3.** States of the Matched Cohorts at Baseline.

| State | Number (%) of patients |  |  |  |
| --- | --- | --- | --- | --- |
|  | Optum CDM Database |  | IQVIA PMTX+ Database |  |
|  | CAS+IMD<br>(n = 1,116) | No mAb treatment<br>(n = 5291) | CAS+IMD<br>(n = 3280) | No mAb treatment<br>(n = 16 284) |
| Alabama | 4 (0.4) | 20 (0.4) | 257 (7.8) | 1285 (7.9) |
| Arkansas | 11 (1) | 50 (0.9) | 86 (2.6) | 430 (2.6) |
| Arizona | 34 (3) | 157 (3) | 31 (0.9) | 155 (1) |
| California | 29 (2.6) | 141 (2.7) | 64 (2) | 315 (1.9) |
| Colorado | 25 (2.2) | 123 (2.3) | 5 (0.2) | 24 (0.1) |
| Connecticut | 9 (0.8) | 45 (0.9) | 18 (0.5) | 89 (0.5) |
| District of Columbia | 2 (0.2) | 7 (0.1) | 1 (0) | 5 (0) |
| Delaware | 1 (0.1) | 1 (0) | 1 (0) | 5 (0) |
| Florida | 170 (15.2) | 818 (15.5) | 175 (5.3) | 859 (5.3) |
| Georgia | 35 (3.1) | 167 (3.2) | 75 (2.3) | 373 (2.3) |
| Iowa | 18 (1.6) | 80 (1.5) | 14 (0.4) | 70 (0.4) |
| Idaho | 2 (0.2) | 10 (0.2) | 28 (0.9) | 131 (0.8) |
| Illinois | 25 (2.2) | 117 (2.2) | 64 (2) | 320 (2) |
| Indiana | 37 (3.3) | 181 (3.4) | 112 (3.4) | 560 (3.4) |
| Kansas | 1 (0.1) | 5 (0.1) | 45 (1.4) | 221 (1.4) |
| Kentucky | 17 (1.5) | 82 (1.5) | 88 (2.7) | 439 (2.7) |
| Louisiana | 51 (4.6) | 242 (4.6) | 279 (8.5) | 1393 (8.6) |
| Massachusetts | 8 (0.7) | 37 (0.7) | 27 (0.8) | 135 (0.8) |
| Maryland | 31 (2.8) | 151 (2.9) | 115 (3.5) | 573 (3.5) |
| Maine | – | – | 2 (0.1) | 10 (0.1) |
| Michigan | 10 (0.9) | 42 (0.8) | 77 (2.3) | 383 (2.4) |
| Minnesota | 37 (3.3) | 185 (3.5) | 119 (3.6) | 592 (3.6) |
| Missouri | 20 (1.8) | 96 (1.8) | 28 (0.9) | 137 (0.8) |
| Mississippi | 19 (1.7) | 85 (1.6) | 35 (1.1) | 175 (1.1) |
| Montana | – | – | 24 (0.7) | 112 (0.7) |
| North Carolina | 49 (4.4) | 231 (4.4) | 130 (4) | 650 (4) |
| North Dakota | 2 (0.2) | 10 (0.2) | 13 (0.4) | 63 (0.4) |
| Nebraska | 10 (0.9) | 43 (0.8) | 21 (0.6) | 104 (0.6) |
| New Hampshire | 1 (0.1) | 1 (0) | 8 (0.2) | 40 (0.2) |
| New Jersey | 34 (3) | 169 (3.2) | 74 (2.3) | 365 (2.2) |
| New Mexico | 4 (0.4) | 18 (0.3) | 19 (0.6) | 95 (0.6) |
| Nevada | – | – | 3 (0.1) | 14 (0.1) |
| New York | 50 (4.5) | 233 (4.4) | 102 (3.1) | 504 (3.1) |
| Ohio | 55 (4.9) | 269 (5.1) | 94 (2.9) | 470 (2.9) |
| Oklahoma | 13 (1.2) | 65 (1.2) | 165 (5) | 820 (5) |
| Pennsylvania | 26 (2.3) | 121 (2.3) | 144 (4.4) | 713 (4.4) |
| Rhode Island | 1 (0.1) | 1 (0) | 58 (1.8) | 275 (1.7) |
| South Carolina | 12 (1.1) | 56 (1.1) | 73 (2.2) | 365 (2.2) |
| South Dakota | 5 (0.4) | 21 (0.4) | 18 (0.5) | 79 (0.5) |
| Tennessee | 20 (1.8) | 98 (1.9) | 38 (1.2) | 190 (1.2) |
| Texas | 151 (13.5) | 750 (14.2) | 464 (14.1) | 2320 (14.2) |
| Utah | 21 (1.9) | 96 (1.8) | 5 (0.2) | 25 (0.2) |
| Virginia | 15 (1.3) | 71 (1.3) | 45 (1.4) | 223 (1.4) |
| Washington | 2 (0.2) | 9 (0.2) | 4 (0.1) | 20 (0.1) |
| Wisconsin | 45 (4) | 181 (3.4) | 22 (0.7) | 110 (0.7) |
| West Virginia | 4 (0.4) | 6 (0.1) | 9 (0.3) | 43 (0.3) |
| Wyoming | — | — | 1 (0) | 5 (0) |

Abbreviations: CAS+IMD, casirivimab and imdevimab; CDM, Clinformatics Data Mart; mAb, monoclonal antibody; PMTX+, Pharmetrics Plus.

